# Evaluation of a Proprietary Fenugreek Extract, on Muscle Strength, Endurance, Total Serum Testosterone and Athletic Performance in Male Subjects during Resistance Training – A Randomized, Double Blind, Placebo Controlled, Comparative, Interventional, Prospective Study

**DOI:** 10.1101/2025.08.13.25333582

**Authors:** Sanjay Tamoli, Narendra Mundhe, Ganesh Kolhe, Kiran Khatau, Ruby Dubey, Ninad N. Mulye

## Abstract

**Background:** Fenugreek is known to improve muscle strength, stamina and serum testosterone level during resistance training.

**Objectives:** A randomized, double blind, placebo controlled, comparative, interventional, prospective clinical study was conducted to evaluate efficacy and safety of a botanical extract of Fenugreek in Male Subjects during resistance training.

**Methods:** Subjects were advised to take given product (FENUDAAT^TM^ or matching placebo) in a dose of 1 capsule (275 mg) twice daily after meals with water for 8 weeks. A total of 43 healthy participants (22 in FENUDAAT^TM^ group and 21 in placebo group) completed the study. Study subjects participated in a supervised 4-day per week resistance-training program for 8 weeks. The efficacy testing outcomes included serum testosterone (total) level, muscle strength and repetitions to failure, metabolic markers for anabolic activity (serum creatinine and blood urea nitrogen), and body fat percentage. Standard safety measurements such as adverse events monitoring, vital signs, hematology and biochemistry were performed.

**Results:** A significant increase in 1 Repeat Measure (RM) Bench Press, 1 RM Leg Press responses and repetitions to failure in bench press and leg press was observed in the FENUDAAT^TM^ group, compared to Placebo group. A significant reduction in weight and BMI was observed in the FENUDAAT^TM^ group, compared to placebo group. A significant increase in total serum testosterone, strength, stamina, energy, and endurance was observed in the FENUDAAT^TM^ group, compared to placebo group. FENUDAAT^TM^ was safe and well tolerated as there were no adverse events reported.

**Conclusion:** The study concludes that regular consumption of the FENUDAAT^TM^ Capsule improves serum testosterone, strength, stamina, energy, and endurance during resistance training.

## Introduction

Athletes, and other individuals who regularly exercise, are consistently seeking out ways to improve physical performance through supplements and other dietary changes. Areas of desired improvement often include strength, stamina, and overall energy. When developing supplements in this space, it is important to follow strict standards of safety, in order to abide by regulatory standards of each athletic organization. There has been a lot of interest in nutritional supplements for muscle building and performance enhancement, as these products may be safer but equally efficacious, when compared to traditional performance enhancing methods. Currently, many traditional herbal medicines and supplements are being studied for their potential benefits of performance enhancement. Extensive scientific studies are required to establish the safe use of these supplements.

Resistance training is a term that implies the use of load, machinery, or one’s own body weight while exercising various muscle groups. It is a common method used to improve the function and performance of muscles and increase muscle mass. [1–2] Other benefits include improved physical performance, movement control, walking speed, functional independence, cognitive abilities, and self-esteem. It may also improve an individual’s overall health status and may potentially be useful in the management of conditions such as diabetes, cardiovascular health conditions, joint and muscle related conditions etc. [3–5]

Currently, many athletes and other regularly exercising individuals use Fenugreek extract (*Trigonella foenum groecum)* in the form of nutritional supplements. In traditional medicine, Fenugreek has been used in the management of conditions such as diabetes, arthritis, and in post-partum conditions in women. It is considered as a GRAS (Generally Recognized as Safe) ingredient by the US Food and Drug Administration. While Fenugreek has been extensively studied for its lipid lowering and anti-inflammatory properties it has also shown potential weight reducing and fat reducing properties. [6,7] Fenugreek has also been reported to enhance endurance capacity and the utilization of fatty acids as an energy source. [8–10]

These effects of Fenugreek have been attributed to its aromatase activity and 5α-reductase inhibition which improves total testosterone levels by blocking its conversion to Estrogen and dihydrotestosterone, respectively. Increased testosterone levels are correlated with increased muscle size and strength in men. Apart from these effects there is also the added benefit of decreasing body weight and fat, in addition to an increase in strength, libido, energy levels, and positive mood. Increased protein synthesis also results in muscles taking up proteins, resulting in increased muscle synthesis and an improvement in strength. Fenugreek seeds are rich in steroidal compounds such as diosgenin, which is a key precursor required for the synthesis of a number of sex hormones, including testosterone. In animal studies, diosgenin has also been shown to augment overall weight, muscle growth and improve glucose metabolism by promoting adipocyte differentiation and inhibiting inflammation in adipose tissues.

Lodaat Pharma has developed FENUDAAT^TM^ a proprietary extract of Fenugreek Seeds (*Trigonella foenum-graecum*). It is a nutritional supplement intended for use by athletes and individuals who regularly exercise for muscle building and improving performance. Based on the activities of Fenugreek, a hypothesis was postulated that fenugreek will be useful in enhancing performance and improving strength and endurance in individuals living a healthy lifestyle. To test the hypothesis, a randomized, double blind, placebo controlled, comparative, clinical study was planned to study the effect of fenugreek given in the form of 275mg capsules twice a day in male subjects undergoing resistance training.

## Materials & Methods

### Settings and locations

The study was conducted at KVTR Ayurveda College and Hospital, Boradi, Dhule District, Maharashtra, India.

### Study design

This was a randomized, double blind, placebo controlled, comparative, interventional, prospective clinical study.

### Ethical considerations

The study was approved by the Institutional Ethics Committee, KVTR Ayurveda College and Hospital, Boradi, Dhule District, India (Letter no. AMB/331/2021-22 dated 22/06/2021) and registered with Clinical Trial Registry - India (CTRI/2021/07/034548, Registered on: 02/07/2021). This clinical trial was conducted in accordance with the Declaration of Helsinki.

#### Enrollment of patients

The study was carried out on healthy male volunteers of age between 18 and 45 years (both inclusive), with normal health status based on clinical and laboratory examination and who consented to participate in the study. The study was carried out and reported adhering to CONSORT statement.

#### Study duration & Visits

The total duration of the study was 8 weeks. The study visits were planned and outcomes were assessed on Screening Visit, Baseline Visit (Day 0), week 4 and week 8 i.e. end of study.

### Eligibility criteria for participants

#### Inclusion Criteria

Healthy male volunteers of age between 18 and 45 years (both inclusive), with normal health status based on clinical and laboratory examination, moderately active (regular aerobic exercise for at least 4 hours per week), with no known musculoskeletal pathology, who were ready to provide written informed consent, and who were ready to willingly participate and follow the protocol requirements of the clinical study were included in the study.

#### Exclusion Criteria

Subjects with known musculoskeletal pathology, using systemic corticosteroids within 2 months of screening, and/or using any other investigational product within 1 month prior to randomization were excluded from the study. Subjects with uncontrolled diabetes mellitus, hypertension, subjects with known tuberculosis, HIV, ischemic heart disease, cancer, kidney failure, subjects with significant abnormal laboratory parameters, and subjects with known hypersensitivity to any of the ingredients of FENUDAAT^TM^ were excluded from the study. Other conditions, which in the opinion of the investigators, could have made subject unsuitable for enrolment or could have interfered with his participation in and completion of the protocol were also excluded from the study.

#### Laboratory Investigations

Laboratory investigations, including CBC, ESR, haemoglobin levels, fasting blood sugar levels, liver function tests, renal function tests, lipid profile, and serum total testosterone were performed at screening visit and at the end of the study.

#### Sample size

The primary population for this study was per protocol population. With over 40 participants, the sample size was considered sufficient for analysis with 80% power, Type I error to be 5% and Type II error to be 20%. A total of 43 participants were randomized in the study, 22 in Study Product Group (FENUDAAT^TM^ Group) and 21 in Control Group (Placebo Group).

#### Randomization

Block randomization with centre as the stratum was done. Subjects meeting eligibility criteria were randomized 1:1 to one of the two groups.

#### Intervention Details

As per computer generated randomization list, subjects were randomized either to a proprietary fenugreek extract supplement (FENUDAAT^TM^) or matching placebo in 1: 1 ratio. Subjects were advised to take given product (FENUDAAT^TM^ or matching placebo) in a dose of 1 capsule (275 mg) twice daily after meals with water for 8 weeks.

#### Primary and secondary outcome measures

Primary outcome measure was comparative change in muscle strength and endurance during resistance training (bench press and leg press) at baseline, week 4, and week 8 (end of study) of the resistance training program between two groups.

Secondary outcome measures were comparative change in skinfold thickness, body fat percentage, body weight, body mass index, lean body mass at baseline, week 4, week 8 (end of study) of the resistance training program between two groups, comparative change in serum creatinine, blood urea nitrogen (BUN) and serum total testosterone at baseline, and week 8 (end of study) of the resistance training program between two groups, comparative change in participant perceived outcome measures like change in strength, stamina, energy, endurance at baseline, week 4, and week 8 (end of study) of the resistance training program between two groups and comparative change in global assessment for overall change by investigator and subject on CGI Scale between two groups at the end of the study.

#### Assessment of Safety

Safety of the study product was assessed by evaluating adverse events, overall safety, and tolerability of the product by the physician and subject on global assessment scale. All adverse events data were listed per subject including severity grading, relationship with investigational product and relationship of the adverse event to other causality, action taken and outcome of the adverse event. Any clinically significant changes in laboratory parameters were also reported.

#### Study methodology

On screening visit (Day -3), a written informed consent was obtained from subjects for their participation in the study. Subject’s demographic details and medical history was recorded. Subject’s general and systemic examinations, including vitals, were performed. Subject’s laboratory investigations i.e., CBC, ESR, haemoglobin levels, fasting blood sugar levels, liver function tests, renal function tests, lipid profile, and serum total testosterone were conducted.

On baseline visit (Day 0), subjects were recruited if they met all the inclusion criteria. Subject’s general and systemic examinations including vitals were performed. Recruited subjects were instructed to refrain from exercise for 48 hrs and fast for 12 hrs before randomization and baseline assessment visit. As per computer generated randomization list, subjects were randomized either to the FENUDAAT^TM^ or matching placebo in 1: 1 ratio. Subjects were advised to take given product (FENUDAAT^TM^ or matching placebo) in a dose of 1 capsule (275 mg) twice daily after meals with water for 8 weeks.

On baseline visit (Day 0), 4 week and end of study visit (Week 8), subject’s skinfold thickness of subcutaneous tissue of abdominals, thigh, triceps, and chest were measured with the help of skinfold caliper by well-trained site personnel. Subject’s body fat percentage and lean body mass was estimated using bioelectrical impedance machine. Subject’s weight and BMI was measured. Participant perceived outcome measures like change in strength, stamina, energy, endurance were assessed on 7-point scale ranging from +3 to −3: −3 (very dissatisfied), −2 (somewhat dissatisfied), −1 (a little dissatisfied), 0 (neutral), +1 (a little satisfied), +2 (somewhat satisfied), and +3 (very satisfied).

#### Resistance training and muscle strength measurement

Briefly, each subject performed 1RM (in weight training, it is the maximum amount of weight that a person can possibly lift for one repetition) lifts on the isotonic bench press. Initially, subjects warmed up (2 sets of 8–10 repetitions at approximately 50% of anticipated maximum) on the bench press and then performed successive 1RM lifts starting at about 70% of anticipated 1RM and increase by 5 kg until they reached 1RM. Subjects were allowed to rest again, and warmed up by performing 2 sets of the bench press at 60% and 80% of the resistance. Then, subjects completed as many repetitions as possible with a resistance of 80% of their 1RM bench press. Subjects were allowed to rest for 10 min and then subjects warmed up on the 45° leg press (2 sets of 8–10 repetitions at approximately 50% of anticipated maximum). The same procedure as that of 1RM bench press was adopted for 1RM leg press. The repetitions to failure in bench press and leg press were recorded from the number of maximum repetitions that subjects could have completed with a resistance of 80% of their 1RM bench/leg press.

On final follow up visit (Week 8), subject’s global evaluation for overall change and Investigator’s global evaluation for overall change were done. All the subjects were closely monitored for any adverse events/ adverse drug reactions from baseline visit till the end of the study. Subject’s laboratory investigations i.e., CBC, ESR, Hb%, BSL-F, Liver function tests, Renal function tests, Lipid profile and Serum total testosterone were done. Subjects were asked to stop study product after completion of 8 weeks and will be advised to take investigator’s advice for further management.

#### Statistical analysis

The data were analysed for central tendencies (mean, median), range, standard error, and standard deviation. Data were tabulated and graphically shown using standard format and MS Excel. Statistical tests were conducted to compare study groups as per the distribution (normality) Student’s T test (normative), Mann-Whitney statistic (non-parametric), Chi-square statistic (categorical), and ANOVA. The level of significance at p < 0.05 (two sided) was considered significant. Both intent-to-treat and per protocol completer analysis were performed when appropriate. Standard statistical software programs were used (GraphPad InStat Version 3.6).

## Results

A total of 43 subjects were screened for inclusion in the study. All the 43 subjects were randomized as there were no screen failures. Out of the 43 randomized subjects, 22 were randomized to the fenugreek extract group and remaining 21 were randomized to the placebo group. Since there were no dropouts, all 43 subjects completed the study. The details are presented in the CONSORT chart.

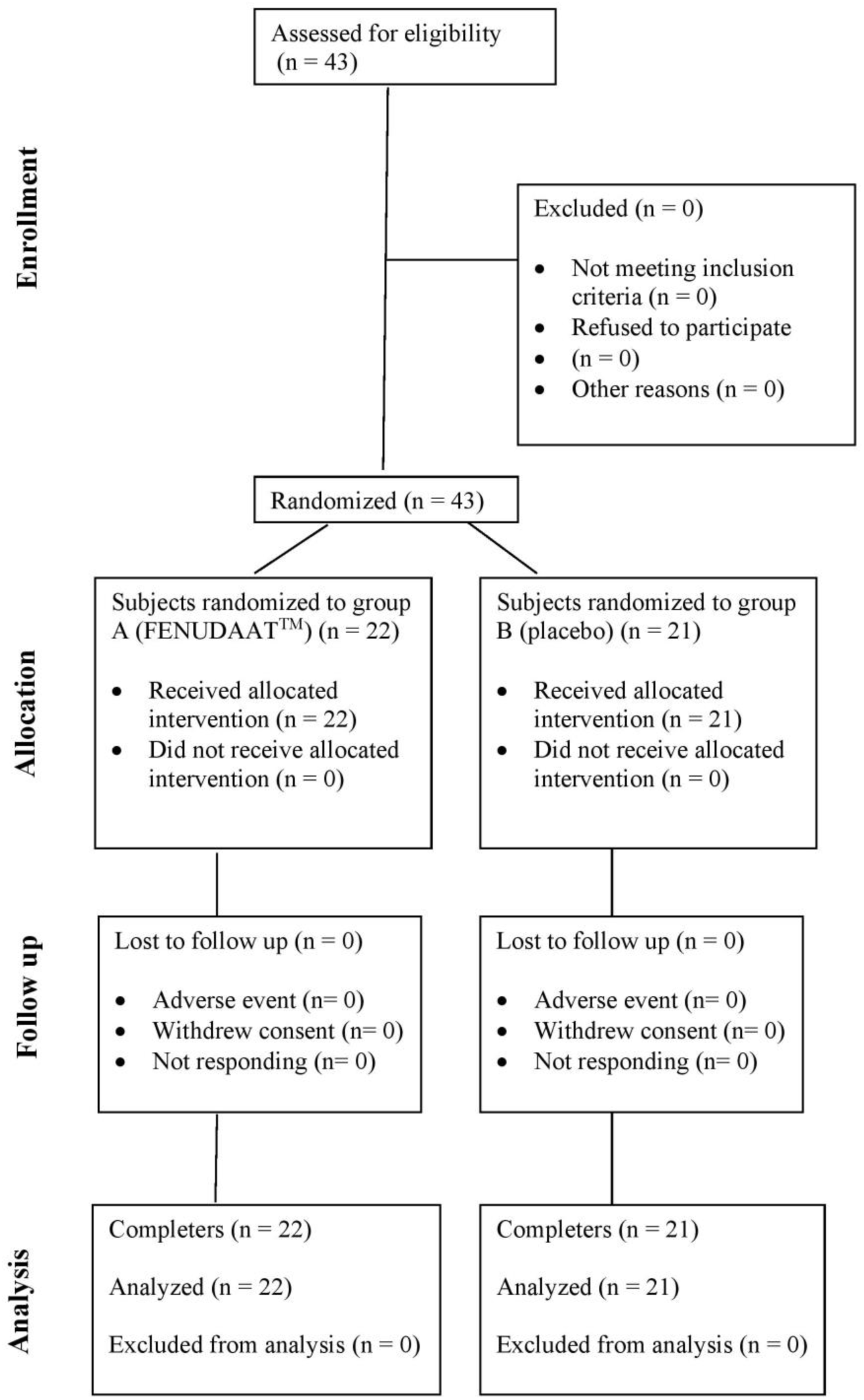

### Baseline demography

Details regarding gender, average age, height, weight and BMI for both groups are presented in Table 1.

**Table 1:** Baseline Demography.

|  | **FENUDAAT™ Group**<br>**(n=22)** | **Placebo Group (n=21)** |
| --- | --- | --- |
| Male (n) | 22 | 21 |
| Age in Years | 21.4±1.14 | 20.6±1.12* |
| Height in Meter | 1.70±0.05 | 1.69±0.05* |
| Weight in Kg | 64.95±10.68 | 63.69±13.63* |
| BMI (Kg/m<sup>2</sup>) | 22.32±3.05 | 22.10±4.68* |

\*Nonsignificant difference (p>0.05)

### Assessment of Primary Outcome

#### Comparative changes in muscle strength and endurance during resistance training

The FENUDAAT^TM^ group showed a statistically significant increase in 1 RM Bench Press and 1 RM Leg Press responses as compared to placebo at the 4-week and 8-week marks. A statistically significant increase in repetitions to failure in bench press and leg press were also observed in FENUDAAT^TM^ group on week 4 and week 8 as compared to placebo. The details are presented in table 2.

**Table 2:** Comparative changes in muscle strength and endurance during resistance training.

|  | **FENUDAAT™ Group n=22** |  |  | **Placebo Group n=21** |  |  | **Between group P-value** |
| --- | --- | --- | --- | --- | --- | --- | --- |
| **Parameters** | **Baseline (n=22)** | **Week 4 (n=22)** | **Week 8 (n=22)** | **Baseline (n=21)** | **Week 4 (n=21)** | **Week 8 (n=21)** |  |
| **1 RM Bench Press (Kg)** | 66.90<br>±14.92 | 71.72<br>±15.86 | 79.90<br>±18.48 | 65.23<br>±11.58 | 65.23<br>±11.58 | 68.19<br>±11.06 | 0.0162 |
|  |  | p<0.0001 | p<0.0001 |  | p=1 | p<0.0001 |  |
| **1 RM Leg Press (Kg)** | 75.06<br>±14.71 | 81.12<br>±16.48 | 91.58<br>±19.86 | 73.41<br>±13.88 | 73.85<br>±13.52 | 76.91<br>±12.92 | 0.0068 |
|  |  | p<0.0001 | p<0.0001 |  | p=0.3293 | p<0.0001 |  |
| **Repetitions to Failure in Bench Press (n)** | 7.00<br>±1.23 | 7.63<br>±1.13 | 8.31<br>±1.24 | 5.95<br>±1.39 | 5.95<br>±1.39 | 6.23<br>±1.13 | <0.0001 |
|  |  | P=0.0039 | P=0.0001 |  | p>0.9999 | P=0.1094 |  |
| Repetitions to Failure in Leg Press (n) | 7.09<br>±1.15 | 7.77<br>±1.23 | 8.36<br>±1.25 | 5.90<br>±1.37 | 6.00<br>±1.30 | 6.28<br>±0.95 | <0.0001 |
|  |  | p=0.0020 | p=0.0002 |  | p=0.5000 | p=0.0557 |  |

### Assessment of Secondary Outcomes

#### Assessment of comparative changes in Skinfold, Thickness, body fat percentage, Body weight, body mass index, lean body mass

There was a statistically significant reduction on parameters including body weight, BMI, skin fold thickness (thighs, triceps and chest) from baseline to 8 weeks with the use of FENUDAAT^TM^ capsules. In the Placebo group, these parameters showed no significant change from baseline to the end of 8 weeks. On analysis between the groups, no significant difference was observed on these parameters.

A significant reduction of lean body mass was observed in the FENUDAAT^TM^ group while a significant increase was observed in the placebo group. Between group analysis however showed non-significant difference. Similarly, body fat percentage showed a significant increase in placebo group which was not observed in FENUDAAT^TM^ group. Between group an analysis did not show significant difference in the two groups. The details are presented in table 3.

**Table 3:** Assessment of comparative changes in Skinfold, Thickness, body fat percentage, Body weight, body mass index, lean body mass.

|  | FENUDAAT™ (Group A) n=22 |  |  | Placebo (Group B) n=21 |  |  | P Value |
| --- | --- | --- | --- | --- | --- | --- | --- |
|  | Baseline (n=22) | Week 4 (n=22) | Week 8 (n=22) | Baseline (n=21) | Week 4 (n=21) | Week 8 (n=21) |  |
| **Weight in Kg** | 64.95<br>±10.68 | 64.54<br>±10.29 | 64.21<br>±10.25 | 63.69<br>±13.63 | 63.79<br>±13.54 | 63.91<br>±13.49 | 0.9350 |
|  |  | p=0.0699 | p=0.0123 |  | p=0.2397 | p=0.0203 |  |
| **BMI (Kg/m<sup>2</sup>)** | 22.32<br>±3.05 | 22.19<br>±2.93 | 22.07<br>±2.89 | 22.10<br>±4.68 | 22.13<br>±4.65 | 22.34<br>±4.56 | 0.8150 |
|  |  | p=0.0935 | p=0.0110 |  | p=0.2385 | p=0.0560 |  |
| **Body Fat percentage (%)** | 18.98<br>±4.49 | 18.65<br>±4.57 | 18.62<br>±4.58 | 16.96<br>±7.50 | 17.11<br>±7.38 | 20.68<br>±10.94 | 0.7409 |
|  |  | p=0.7159 | p=0.2270 |  | p=0.1055 | p=0.0002 |  |
| **Lean Body Mass (%)** | 36.21<br>±2.99 | 36.73<br>±3.18 | 37.83<br>±3.36 | 36.23<br>±4.63 | 36.43<br>± 4.79 | 36.85<br>±4.83 | 0.4453 |
|  |  | p=0.0250 | p<0.0001 |  | p=0.0637 | p=0.0121 |  |
| **Skinfold Thickness** | 9.86<br>±3.15 | 9.36<br>±2.83 | 8.5<br>±2.44 | 9.57<br>±2.90 | 9.38<br>±3.10 | 9.38<br>±3.05 | 0.3017 |
| **Thighs (mm)** |  | p=0.0611 | p=0.0004 |  | p=0.2500 | p=0.3125 |  |
| **Skinfold Thickness** | 9.81<br>±3.69 | 9.13<br>±3.57 | 8.31<br>±3.22 | 10.09<br>±3.33 | 10.19<br>±3.54 | 10.09<br>±3.34 | 0.0669 |
| **Triceps (mm)** |  | p=0.0039 | p=0.0001 |  | p=0.4939 | p>0.9999 |  |
| **Skinfold Thickness** | 7.5<br>±2.82 | 6.68<br>±2.60 | 6.27<br>±2.69 | 6.52<br>±2.15 | 6.57<br>±2.08 | 6.71<br>±2.17 | 0.5584 |
| **Chest (mm)** |  | p=0.0029 | p=0.0010 |  | p=0.6657 | p=0.1623 |  |

#### Assessment of comparative change in Serum Total Testosterone Parameters in two groups

There was a significant increase in the serum Testosterone levels from baseline to the end of 8 weeks with the use of FENUDAAT^TM^ while the levels decreased in placebo group. However, analysis between the group showed non-significant change. Refer Table 4.

**Table 4:** Assessment of comparative change in Serum Total Testosterone Parameters.

| Test parameter | FENUDAAT™ (Group A) n=22 |  | Placebo (Group B) n=21 |  | p value between groups |
| --- | --- | --- | --- | --- | --- |
|  | Baseline | 8 Weeks | Baseline | 8 Weeks |  |
| **Serum Total Testosterone (ng/dL)** | 474.66<br>±228.23 | 515.23<br>±224.89 | 481.94<br>±292.05 | 453.74<br>±242.52 | 0.2317 |
|  |  | p<0.001 |  | P=0.516 |  |

#### Assessment of comparative change in Blood Urea Nitrogen (BUN) in the two groups

There were no significant changes from baseline to 8 weeks on Sr. Creatinine levels in both the FENUDAAT^TM^ group and Placebo Group and in a between group analysis. The levels were within normal range in both the groups at both time points. Sr. BUN levels showed significant increase in the FENUDAAT^TM^ group while there was nonsignificant change in placebo group. Between group analysis showed no significant difference. The levels were within normal range at both time points in both study groups.

**Table 4:** Assessment of comparative change in Sr. Creatinine, Blood Urea Nitrogen (BUN)

| Parameter | FENUDAAT™ Group (n=22) |  | Placebo Group (n=21) |  | P value between groups |
| --- | --- | --- | --- | --- | --- |
|  | Baseline | 8 Weeks | Baseline | 8 Weeks |  |
| **Sr.**<br>**Creatinine**<br>**(mg/dL)** | 0.67±0.17 | 0.73±0.09 | 0.68±0.15 | 0.71±0.09 | 0.4282 |
|  |  | p=0.0612 |  | p=0.7620 |  |
| **BUN**<br>**(mg/dL)** | 14.71±2.70 | 16.77±3.06 | 14.86±2.80 | 15.57±2.92 | 0.1389 |
|  |  | p=0.0016 |  | p=0.2356 |  |

#### Assessment of Changes in Participant perceived outcome measure (i.e., changes in Strength, Stamina, Energy, Endurance)

In the FENUDAAT^TM^ group, a significant improvement in strength, stamina, energy, and endurance was observed on week 4 and week 8. A significant improvement in strength, stamina, energy, and endurance was observed only at week 8 in placebo group. Between group comparison demonstrated a significant improvement in FENUDAAT^TM^ group compared to placebo group. The details are presented in Table 5 and Graph 1.

**Table 5:** Assessment of comparative changes in Strength stamina, energy, and endurance between two groups.

| **Parameter** | **Drug** | **Baseline** | **Week 4** | **Week 8** |
| --- | --- | --- | --- | --- |
| **Energy** | **FENUDAAT** | 38.64 | 58.33 | 79.54 |
|  | **Placebo** | 38.1 | 38.89 | 43.65 |
| **Strength** | **FENUDAAT** | 36.5 | 50.55 | 65.2 |
|  | **Placebo** | 35.21 | 38.25 | 42.11 |
| **Stamina** | **FENUDAAT** | 39.9 | 55.22 | 75.56 |
|  | **Placebo** | 36.9 | 40.21 | 41.53 |
| **Endurance** | **FENUDAAT** | 40.15 | 60.22 | 75.59 |
|  | **Placebo** | 39.22 | 40.05 | 41.8 |

**Graph 1:**
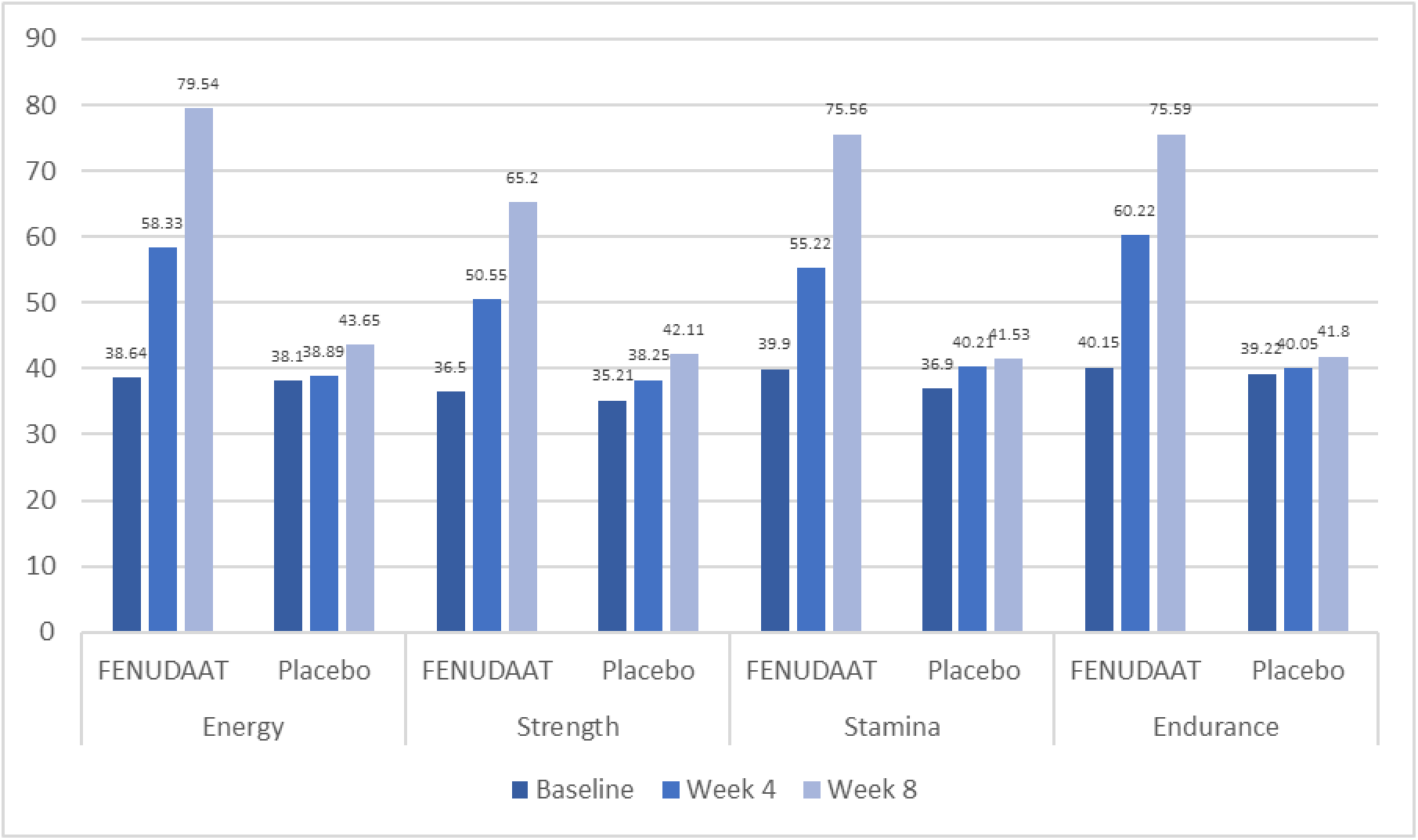
Comparative Assessment of Changes in Strength, Stamina, Energy, and Endurance Between Two Groups.

#### Global assessment of overall change assessed by investigator and by subjects

In the FENUDAAT^TM^ group, as per investigators and subject’s assessment, 16 (72.73%) subjects reported minimal to very much improvement while 6 (27.27%) subjects reported no change in their condition. In the placebo group, as per investigators and subject’s assessment, 3 (14.29%) subjects reported minimal improvement while 18 (85.71%) subjects reported no change in their condition. When compared between group, the difference was statistically significant. The details are presented in table 5.

#### Safety assessment by evaluation of occurrence of AE

At the end of the study no significant change in any of the vital parameters such as pulse rate, respiratory rate, temperature, systolic and diastolic blood pressure within and between the groups was observed. No adverse events were reported by any subjects of both the groups. All the subjects well tolerated study products.

### Safety Lab Parameters

#### Assessment of CBC Parameters in two groups

Assessment of safety related laboratory parameters like CBC, liver function tests and fasting blood sugar levels showed no significant difference between the two groups from baseline to 8 weeks. These levels were observed to be within normal range at both baseline and 8 weeks. Details in Table 6, 7 and 8.

**Table 6:**
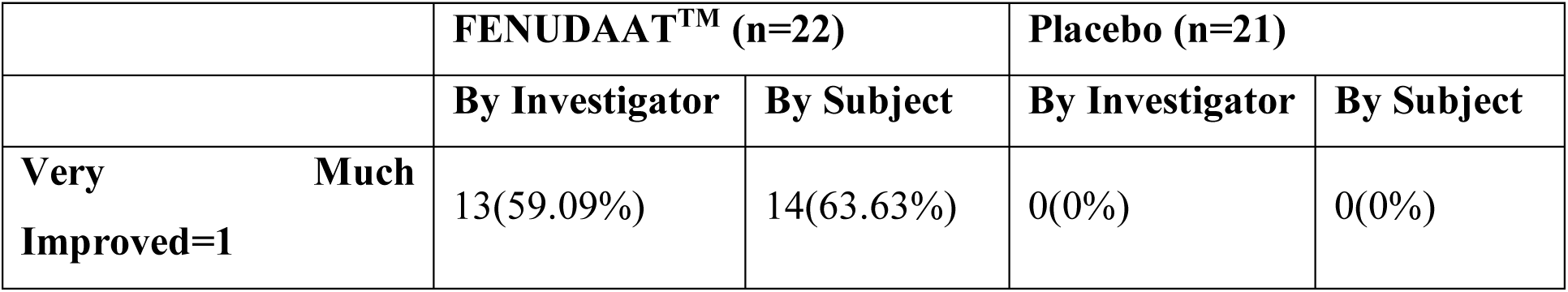
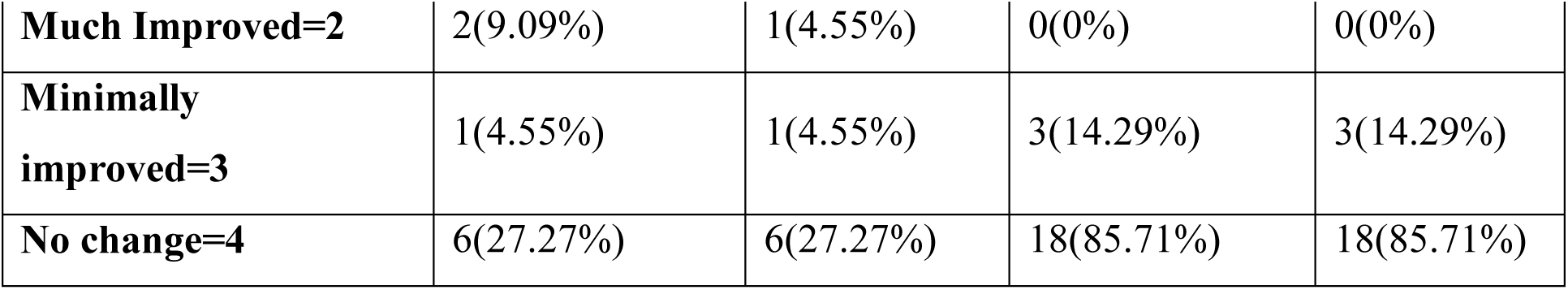
Global assessment of overall change assessed by investigator and by subjects.

|  | FENUDAAT™ (n=22) |  | Placebo (n=21) |  |
| --- | --- | --- | --- | --- |
|  | By Investigator | By Subject | By Investigator | By Subject |
| **Very Much Improved=1** | 13(59.09%) | 14(63.63%) | 0(0%) | 0(0%) |
| **Much Improved=2** | 2(9.09%) | 1(4.55%) | 0(0%) | 0(0%) |
| **Minimally improved=3** | 1(4.55%) | 1(4.55%) | 3(14.29%) | 3(14.29%) |
| **No change=4** | 6(27.27%) | 6(27.27%) | 18(85.71%) | 18(85.71%) |

**Table 7:** Assessment of CBC Parameters in two groups.

| Parameter | FENUDAAT™ Group |  | Placebo Group |  | P value between groups |
| --- | --- | --- | --- | --- | --- |
|  | Baseline | 8 Weeks | Baseline | 8 Weeks |  |
| **Hb% (g/dL)** | 14.09<br>±0.27 | 14.91<br>±1.84 | 14.00<br>±0.62 | 14.40<br>±1.13 | 0.2876 |
|  |  | p=0.0025 |  | p=0.0193 |  |
| **RBC (Million/μL)** | 4.35<br>±1.04 | 4.41±1.06 | 4.45±1.13 | 4.86±1.78 | 0.9183 |
|  |  | p=0.4196 |  | p=0.7916 |  |
| **WBC (Cells/ μL)** | 7042.72<br>±1600.17 | 7572.72<br>±1093.78 | 7209.52<br>±1345.69 | 7309.52<br>±994.43 | 0.1372 |
|  |  | p=0.0089 |  | p=0.6514 |  |
| **Platelets (100,000/uL)** | 2.52<br>±0.58 | 2.91<br>±0.51 | 2.78<br>±0.52 | 3.03<br>±0.45 | 0.4228 |
|  |  | p<0.0001 |  | p=0.0085 |  |
| **ESR** | 8.50<br>±0.91 | 7.58<br>±1.67 | 8.19<br>±0.74 | 7.57<br>±0.59 | 0.1773 |
|  |  | p=0.0028 |  | p=0.0112 |  |

**Table 8:**
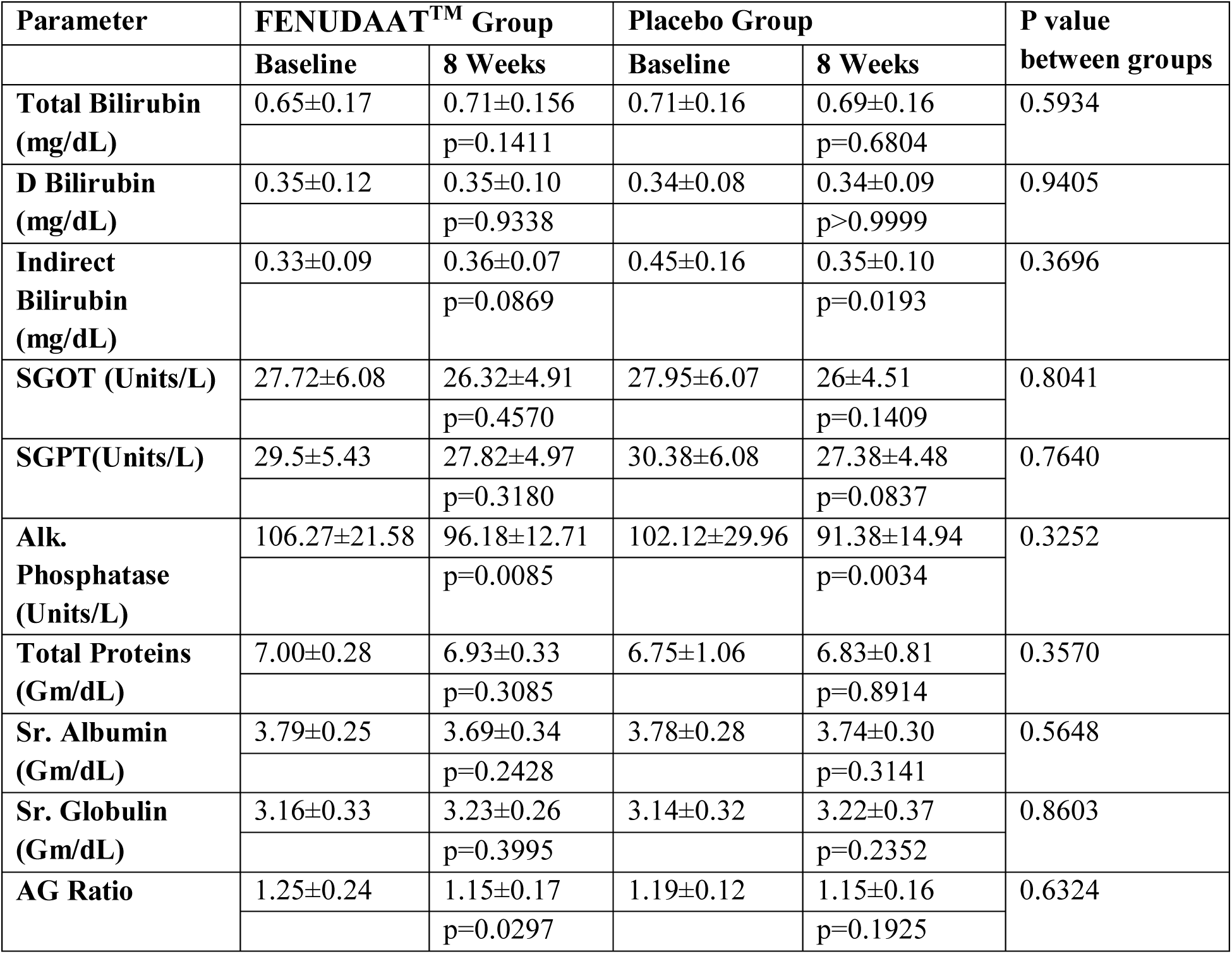
Assessment of Liver function tests in two groups.

| Parameter | FENUDAAT™ Group |  | Placebo Group |  | P value between groups |
| --- | --- | --- | --- | --- | --- |
|  | Baseline | 8 Weeks | Baseline | 8 Weeks |  |
| Total Bilirubin (mg/dL) | 0.65±0.17 | 0.71±0.156 | 0.71±0.16 | 0.69±0.16 | 0.5934 |
|  |  | p=0.1411 |  | p=0.6804 |  |
| D Bilirubin (mg/dL) | 0.35±0.12 | 0.35±0.10 | 0.34±0.08 | 0.34±0.09 | 0.9405 |
|  |  | p=0.9338 |  | p>0.9999 |  |
| Indirect Bilirubin (mg/dL) | 0.33±0.09 | 0.36±0.07 | 0.45±0.16 | 0.35±0.10 | 0.3696 |
|  |  | p=0.0869 |  | p=0.0193 |  |
| SGOT (Units/L) | 27.72±6.08 | 26.32±4.91 | 27.95±6.07 | 26±4.51 | 0.8041 |
|  |  | p=0.4570 |  | p=0.1409 |  |
| SGPT(Units/L) | 29.5±5.43 | 27.82±4.97 | 30.38±6.08 | 27.38±4.48 | 0.7640 |
|  |  | p=0.3180 |  | p=0.0837 |  |
| Alk. Phosphatase (Units/L) | 106.27±21.58 | 96.18±12.71 | 102.12±29.96 | 91.38±14.94 | 0.3252 |
|  |  | p=0.0085 |  | p=0.0034 |  |
| Total Proteins (Gm/dL) | 7.00±0.28 | 6.93±0.33 | 6.75±1.06 | 6.83±0.81 | 0.3570 |
|  |  | p=0.3085 |  | p=0.8914 |  |
| Sr. Albumin (Gm/dL) | 3.79±0.25 | 3.69±0.34 | 3.78±0.28 | 3.74±0.30 | 0.5648 |
|  |  | p=0.2428 |  | p=0.3141 |  |
| Sr. Globulin (Gm/dL) | 3.16±0.33 | 3.23±0.26 | 3.14±0.32 | 3.22±0.37 | 0.8603 |
|  |  | p=0.3995 |  | p=0.2352 |  |
| AG Ratio | 1.25±0.24 | 1.15±0.17 | 1.19±0.12 | 1.15±0.16 | 0.6324 |
|  |  | p=0.0297 |  | p=0.1925 |  |

#### Assessment of Lipid Profile Parameters in two groups

In the FENUDAAT^TM^ group, a significant reduction in Sr. Cholesterol, Sr. Triglycerides, TC: HDL and LDL: HDL levels were observed from baseline to 8 weeks while a significant increase was observed in HDL cholesterol levels. In the placebo group, a significant increase in Sr. Cholesterol, Sr. Triglycerides, HDL and VLDL levels was observed. When compared between the groups, no significant difference was observed. The details are presented in table 9.

**Table 9:**
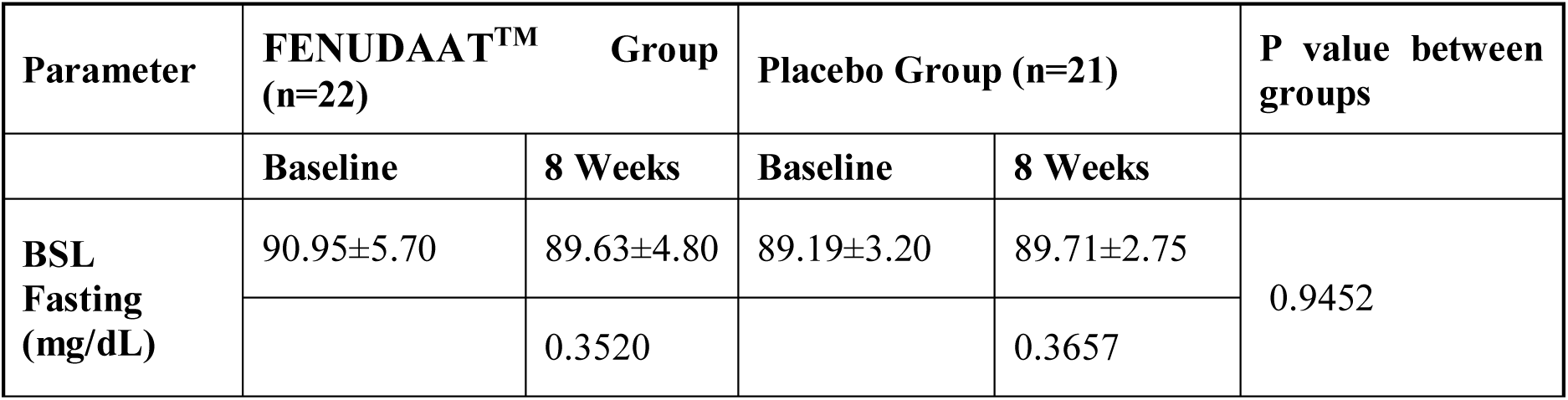
Assessment of change in BSL-Fasting in two groups.

| Parameter | FENUDAAT™ Group (n=22) |  | Placebo Group (n=21) |  | P value between groups |
| --- | --- | --- | --- | --- | --- |
|  | Baseline | 8 Weeks | Baseline | 8 Weeks |  |
| BSL Fasting (mg/dL) | 90.95±5.70 | 89.63±4.80 | 89.19±3.20 | 89.71±2.75 | 0.9452 |
|  |  | 0.3520 |  | 0.3657 |  |

**Table 10:** Assessment of Lipid Profile Parameters in two groups.

| Parameters | FENUDAAT™ Group |  | Placebo Group |  | P value between groups |
| --- | --- | --- | --- | --- | --- |
|  | Baseline | 8 Weeks | Baseline | 8 Weeks |  |
| Sr. Cholesterol (mg/dL) | 177.59±22.50 | 166.95±18.57 | 167.52±16.30 | 169.66±16.98 | 0.6205 |
|  |  | p=0.0011 |  | p=0.0103 |  |
| Sr. Triglycerides (mg/dL) | 129.86±36.99 | 119.27±33.93 | 123.62±30.32 | 126.19±31.44 | 0.3753 |
|  |  | p=0.0003 |  | p=0.0012 |  |
| HDL (mg/dL) | 39.09±4.70 | 44.09±6.79 | 39.90±4.83 | 42.52±5.47 | 0.4112 |
|  |  | p<0.0001 |  | p=0.0004 |  |
| LDL (mg/dL) | 105.86±21.68 | 95.86±14.65 | 97.14±11.31 | 97.95±11.64 | 0.3127 |
|  |  | p=0.0001 |  | p=0.4717 |  |
| VLDL (mg/dL) | 27.11±10.50 | 25.42±8.86 | 23.71±4.61 | 28±9.03 | 0.3555 |
|  |  | p=0.9923 |  | p=0.0056 |  |
| TC: HDL | 4.42±0.87 | 3.75±0.60 | 4.36±1.27 | 4.03±0.56 | 0.1252 |
|  |  | p=0.0002 |  | p=0.1309 |  |
| LDL: HDL | 2.95±1.19 | 2.26±0.45 | 2.47±0.46 | 2.33±0.36 | 0.6187 |
|  |  | p=0.0073 |  | p=0.0005 |  |

## Discussion

The present study was conducted to demonstrate the androgenic and anabolic effects of the FENUDAAT^TM^ capsule containing 275 mg of fenugreek seeds extract FENUDAAT^TM^ (*Trigonella foenum-graecum*) in male subjects during resistance training. The results of the study demonstrated a statistically significant increase in 1 RM Bench Press and 1 RM Leg Press responses in FENUDAAT^TM^ group compared placebo. In addition, an 8-week supplementation of FENUDAAT^TM^ capsule resulted in a statistically significant increase in repetitions to failure in bench press and leg press as compared to placebo.

A statistically significant reduction in weight and BMI was observed in the FENUDAAT^TM^ group. In contrast, a statistically significant increase in weight was observed in placebo group. A statistically significant reduction in skin fold thickness of the thighs, triceps and chest was observed in the FENUDAAT^TM^ group, whereas in the placebo group, no statistically significant difference was observed. A significant increase in total serum testosterone was observed in the FENUDAAT^TM^ group. In the Placebo group no notable change in total serum testosterone was observed. However, the increase in the FENUDAAT^TM^ group was non-significant as compared to placebo in a between-group analysis. A significant improvement in strength, stamina, energy, and endurance was observed as compared to placebo over a period of 4 weeks and 8 weeks during use of the FENUDAAT^TM^ capsules. The results of the present study are in line with the earlier studies conducted where FENUDAAT^TM^ ation demonstrated androgenic and anabolic effects in male subjects during resistance training. [11]

Fenugreek contains glycosides including saponins, sapogenins (e.g., diosgenin), similagenin, savsalpogenin, and yuccagenin [12,13]. A statistically significant increase in serum total testosterone after 8 weeks supplementation of Fenugreek extract has been attributed to its aromatase and 5α-reductase inhibition activity that leads to improvement in total testosterone levels by blocking its conversion to estrogen and dihydrotestosterone, respectively. [14]

An 8-week supplementation of FENUDAAT^TM^ capsules has resulted in a significant increase in 1 RM Bench Press and 1 RM Leg Press responses and a significant increase in repetitions to failure in bench press and leg press. The reason behind this could be the positive effects of the Fenugreek extract on testosterone and muscle strength. Apart from these, there was the added benefit of decreased body weight and body fat. This could be attributed to furostanol glycosides as they have demonstrated potent anabolic and fat burning activity in immature castrated male rats.[15] Also Diosgenin and saponin are known to reduce fat by inhibiting cholesterol absorption, increasing biliary cholesterol secretion and faecal excretion of neutral sterols, thus reducing the liver cholesterol concentrations. Diosgenin also helps to improve glucose metabolism by promoting adipocyte differentiation and inhibiting inflammation in adipose tissues. [16–18]. Increase in testosterone concentrations results in escalated delivery and use by muscle cells. This enhances protein synthesis, causes muscles to take up proteins and results in improved muscle synthesis and strength.[19] Diosgenin is also a key precursor for the synthesis of multiple sex hormones including testosterone. The resulting increase in testosterone may potentially improve mood and libido. [20]

Fenugreek extract is certified as a GRAS ingredient in USA. Therefore, the risk of inherent toxicity is exceptionally low. It is also evident from the present study that by the end of the study no significant change in any of the vital parameters such as pulse rate, respiratory rate, temperature, systolic and diastolic blood pressure was observed. No adverse events were reported by any subjects of both the groups. All the monitored laboratory parameters were within normal limits both at baseline and at the end of the study, suggesting safety of the proposed FENUDAAT^TM^ capsule. One additional benefit of FENUDAAT^TM^ ation observed in the study was its role in reducing the levels of serum cholesterol, triglycerides, low density lipoproteins (LDL) and increasing the levels of high density lipoproteins (HDL). Further clinical studies are warranted to gain a more robust understanding of the effects of fenugreek extract supplementation, particularly in different populations.

## Conclusion

It can be concluded from the results of the study that 8 weeks supplementation with FENUDAAT^TM^ exhibited improvements in muscle strength, stamina, energy, endurance, and a reduction in body weight, fat, and lipid levels. It also demonstrated a significant increase in serum total testosterone, which may support improvements in muscle mass, strength, and positive mood in male subjects during resistance training.

## Funding

The study was funded by Lodaat Pharma, 1415 West 22nd Street-Tower Floor, Oak Brook, Illinois 60523 both financially and materially.

## Authors’ Contributions

All authors are responsible for the content and writing of the paper. All authors have critically contributed to the conceptualization, data collection, analysis, and writing of the paper.

## Institutional Ethical Committee Statement

The study was conducted according to the guidelines of the Declaration of Helsinki, and approved by the Institutional, KVTR Ayurveda College and Hospital, Boradi, Dhule District, India. (Letter no. AMB/331/2021-22 dated 22/06/2021)

## Informed Consent Statement

A written informed consent was obtained from all subjects involved in the study.

## Data Availability Statement

The datasets generated and/or analyzed during the current study are available from the corresponding author on reasonable request.

## Acknowledgments

Authors greatly acknowledge the invaluable collaborative efforts of the study participants. Authors express their gratitude to Lodaat Pharma for sponsoring this study. Authors also thank team at Target Institute of Medical Education and Research, Mumbai, India and Lodaat Pharma, Illinois, US for their priceless support.

## Conflicts of Interest

The authors declare that they have no financial, personal, or professional conflicts of interest that could have influenced the content or outcomes of this manuscript.

## Notes

### Competing Interest Statement

The authors have declared no competing interest.

### Clinical Trial

CTRI/2021/07/034548

## References

1. Stojiljković, N.; Ignjatović, A.; Savić, Z.; Marković, Ž.; Milanović, S. History of resistance training. Adv. Phys. Educ. 2013, 3, 15–22.

2. Kraemer, W. J.; Ratamess, N. A. Fundamentals of resistance training: progression and exercise prescription. Med. Sci. Sports Exerc. 2004, 36, 674–688.

3. Westcott, W. L. Resistance training is medicine: effects of strength training on health. Curr. Sports Med. Rep. 2012, 11, 209–216.

4. Drenowatz, C.; Greier, K. Resistance training in youth – benefits and characteristics. J. Biomed. 2018, 3, 32–39.

5. Md Japilus, S. J.; Hashim, N. H.; Musa, R. N. The effects of exercise order during resistance training on muscular strength. J. Phys. Conf. Ser. 2020, 1529, 022024.

6. Basch, E.; Ulbricht, C.; Kuo, G.; Szapary, P.; Smith, M. Therapeutic applications of fenugreek. Altern. Med. Rev. 2003, 8, 20–27.

7. Ikeuchi, M.; Yamaguchi, K.; Koyama, T.; Sono, Y.; Yazawa, K. Effects of fenugreek seeds (Trigonella foenum-graecum) extract on endurance capacity in mice. J. Nutr. Sci. Vitaminol. (Tokyo) 2006, 52, 287–292.

8. Bhasin, S.; Storer, T. W.; Berman, N.; Callegari, C.; Clevenger, B.; Phillips, J., et al. The effects of supraphysiologic doses of testosterone on muscle size and strength in normal men. N. Engl. J. Med. 1996, 335, 1–7.

9. Tan, R. S.; Culberson, J. W. An integrative review on current evidence of testosterone replacement therapy for the andropause. Maturitas 2003, 45, 15–27.

10. Brodsky, I.; Balagopal, P.; Nair, K. S. Effects of testosterone replacement on muscle mass and muscle protein synthesis in hypogonadal men—a clinical research center study. J. Clin. Endocrinol. Metab. 1996, 81, 3469–3475.

11. Wankhede, S.; Mohan, V.; Thakurdesai, P. Beneficial effects of fenugreek glycoside supplementation in male subjects during resistance training: a randomized controlled pilot study. J. Sport Health Sci. 2016, 5, 176–182.

12. Yadav, S. K.; Sehgal, S. Effect of home processing and storage on ascorbic acid and beta-carotene content of Bathua (Chenopodium album) and fenugreek (Trigonella foenum-graecum) leaves. Plant Foods Hum. Nutr. 1997, 50, 239–247.

13. Billaud, C.; Adrian, J. Fenugreek: composition, nutritional value and physiological properties. Sci. Aliments 2001, 21, 3–26.

14. Wilborn, C.; Taylor, L.; Poole, C.; Foster, C.; Willoughby, D.; Kreider, R. Effects of a purported aromatase and 5α-reductase inhibitor on hormone profiles in college-age men. Int. J. Sport Nutr. Exerc. Metab. 2010, 20, 457–465.

15. Aswar, U.; Bodhankar, S. L.; Mohan, V.; Thakurdesai, P. A. Effect of furostanol glycosides from Trigonella foenum-graecum on the reproductive system of male albino rats. Phytother. Res. 2010, 24, 1482–1488.

16. Uchida, K.; Takase, H.; Nomura, Y.; Takeda, K.; Takeuchi, N.; Ishikawa, Y. Changes in biliary and fecal bile acids in mice after treatments with diosgenin and β-sitosterol. J. Lipid Res. 1984, 25, 236–245.

17. Somani, B.; Khan, S.; Donat, R. Screening for metabolic syndrome and testosterone deficiency in patients with erectile dysfunction: results from the first UK prospective study. BJU Int. 2010, 106, 688–690.

18. Griggs, R. C.; Kingston, W.; Jozefowicz, R. F.; Herr, B. E.; Forbes, G.; Halliday, D. Effect of testosterone on muscle mass and muscle protein synthesis. J. Appl. Physiol. 1989, 66, 498–503.

19. Kiss, R.; Kovács, T.; Kőszegi, T.; Sárközi, K.; Pázmány, L.; Rácz, K., et al. Diosgenin and its fenugreek based biological matrix affect insulin resistance and anabolic hormones in a rat based insulin resistance model. Biomed. Res. Int. 2019, 2019, 7213913.

20. Aradhana; Rao, A. R.; Kale, R. K. Diosgenin—a growth stimulator of mammary gland of ovariectomized mouse. Indian J. Exp. Biol. 1992, 30, 367–370.

